## supplementary_appendix for "Risk of Coronavirus Disease 2019 (COVID-19) among Those Up-to-Date and Not Up-to-Date on COVID-19 Vaccination by US CDC Criteria"

This supplement has been provided by the authors to give readers additional information about the work.

### Supplementary tables

#### Supplementary Table 1

Supplementary Table 1. Adjusted Associations With Time to COVID-19 When Those Above Age 65 Needed Two Vaccine Doses to Be Considered “Up-To-Date”

| Variables | Adjusted HR (95% CI)^a^ | *P* |
| --- | --- | --- |
| Vaccination status “up-to-date”^b^ | 1.12 (0.95-1.32) | 0.18 |
| Propensity to get tested for COVID-19^c^ | 1.09 (1.08-1.10) | <0.001 |
| Age | 0.997 (0.993-1.001) | 0.15 |
| Male sex | 0.80 (0.70-0.90) | <0.001 |
| Most recent prior SARS-CoV-2 infection^d^ |  |  |
| During Pre-Omicron phase | 1.07 (0.92-1.24) | 0.38 |
| During Omicron BA.1/BA.2 dominant phase | 0.83 (0.72-0.95) | 0.006 |
| During Omicron BA.4/BA.5 dominant phase | 0.27 (0.21-0.34) | <0.001 |
| During Omicron BQ dominant phase | 0.09 (0.04-0.21) | <0.001 |
| Number of prior vaccine doses^e^ |  |  |
| 1 or 2 | 1.71 (1.39-2.12) | <0.001 |
| 3 | 2.13 (1.72-2.63) | <0.001 |
| >3 | 2.15(1.66-2.78) | <0.001 |

Abbreviation: COVID-19, Coronavirus Disease 2019; HR, hazard ratio; CI, confidence interval; SARS-CoV-2, Severe Acute Respiratory Syndrome Coronavirus-2

^a^From a multivariable Cox-proportional hazards regression model.

^b^Time-dependent covariate

^c^Calculated as number of COVID-19 nucleic acid amplification tests done per year of employment at Cleveland Clinic during the course of the pandemic.

^d^Reference: no documented prior infection.

^e^Reference: Zero doses

#### Supplementary Table 2

Supplementary Table 1. Adjusted Associations With Time to COVID-19, When a Person Was Considered “Up-To-Date” Only After 7 Days Had Passed Since Receipt of the Bivalent Vaccine

| Variables | Adjusted HR (95% CI)^a^ | *P* |
| --- | --- | --- |
| Vaccination status “up-to-date”^b^ | 1.04 (0.87-1.24) | 0.70 |
| Propensity to get tested for COVID-19^c^ | 1.09 (1.08-1.10) | <0.001 |
| Age | 0.997 (0.993-1.001) | 0.09 |
| Male sex | 0.80 (0.70-0.90) | <0.001 |
| Most recent prior SARS-CoV-2 infection^d^ |  |  |
| During Pre-Omicron phase | 1.07 (0.92-1.24) | 0.38 |
| During Omicron BA.1/BA.2 dominant phase | 0.83 (0.72-0.95) | 0.006 |
| During Omicron BA.4/BA.5 dominant phase | 0.27 (0.21-0.34) | <0.001 |
| During Omicron BQ dominant phase | 0.09 (0.04-0.21) | <0.001 |
| Number of prior vaccine doses^e^ |  |  |
| 1 or 2 | 1.72 (1.39-2.13) | <0.001 |
| 3 | 2.15 (1.74-2.66) | <0.001 |
| >3 | 2.29 (1.75-2.99) | <0.001 |

Abbreviation: COVID-19, Coronavirus Disease 2019; HR, hazard ratio; CI, confidence interval; SARS-CoV-2, Severe Acute Respiratory Syndrome Coronavirus-2

^a^From a multivariable Cox-proportional hazards regression model.

^b^Time-dependent covariate

^c^Calculated as number of COVID-19 nucleic acid amplification tests done per year of employment at Cleveland Clinic during the course of the pandemic.

^d^Reference: no documented prior infection.

^e^Reference: Zero doses
